# The HEART-GP strategy for ruling out acute coronary syndrome in out-of-hours primary care: a diagnostic accuracy trial protocol

**DOI:** 10.1101/2025.11.27.25341146

**Authors:** Indra M.B. Melessen, Jelle C.L. Himmelreich, Amy Manten, Edanur Sert, Simone van den Bulk, Tobias N. Bonten, Martijn H. Rutten, Eric P. Moll van Charante, Ralf E. Harskamp

**Affiliations:** Amsterdam UMC, University of Amsterdam, Academic Medical Center, Department of General Practice, Meibergdreef 9, 1105 AZ Amsterdam, The Netherlands; Amsterdam Cardiovascular Sciences, Atherosclerosis & Ischemic syndromes, Amsterdam, the Netherlands; Amsterdam Public Health, Personalized Medicine, Amsterdam, The Netherlands; Leiden University Medical Center, Department of Public Health and Primary Care, Albinusdreef 2, 2333 ZA Leiden, The Netherlands; Coöperatie Cohesie, Huisartsenspoedpost Noord-Limburg, Prof. Gelissensingel 20, 5912JX Venlo, the Netherlands; Amsterdam UMC, University of Amsterdam, Academic Medical Center, Department of Public and Occupational Health, Amsterdam Public Health

**Keywords:** Chest pain, primary health care, major adverse cardiac events, hs-troponin POCT, risk stratification scores, after hours care

## Abstract

**Background:** Acute coronary syndrome (ACS) is a life-threatening condition that must be rapidly identified to prevent morbidity and mortality. Differentiating ACS from benign conditions remains difficult in primary care settings due to non-specific and overlapping symptom profiles, and limited diagnostic resources. Current guidelines promote a low threshold for emergency department (ED) referral, resulting in referral rates of 40–70%. Despite this cautious approach, ACS cases are still missed, sometimes with serious consequences. The arrival of high-sensitivity troponin (hs-troponin) point-of-care testing (POCT) may enable safer, faster, and more efficient diagnosis in primary care settings.

**Methods and analysis:** This multicenter prospective diagnostic accuracy trial evaluates the *HEART-GP strategy*, combining a single fingerstick hs-troponin test with clinical assessment and optional ECG. Adults (≥18 years) presenting with acute chest pain or discomfort to one of four participating Dutch OOH-PC centers are eligible. The primary outcome is the occurrence of major adverse cardiovascular events a composite of death, ACS, or urgent revascularization within six weeks. Diagnostic safety (sensitivity, negative predictive value) and efficiency (ED referral reduction) will be compared against standard care. Secondary analyses will assess the value of sex-specific cut-offs and integration with existing risk scores (HEART, INTERCHEST, Marburg Heart Score).

**Anticipated results:** We anticipate that the HEART-GP strategy will demonstrate high diagnostic safety along with marked efficiency gains, with potential for reducing unnecessary ED referrals. If hs-POCT proves reliable, it could be implemented widely to improve the safety and efficiency of acute chest pain evaluation in pre-hospital care.

**Ethics and dissemination:** The study was approved by the CCMO (CIV-NL-22-11-041231, 11-03-2022) and the Medical Research Ethics Committee (MREC) of the Amsterdam UMC (NL82428.000.22, 02-03-2023) and registered in the ISRCTN registry (ISRCTN11954040). Results will be disseminated through peer-reviewed journals, academic conferences, and stakeholder engagement to inform future diagnostic pathways in primary care.

**Strengths and limitations of this study:** *Strengths:*

- An innovative, pragmatic strategy for rapid rule-out of ACS
- High external validity due to prospective, multi-center, real-world OOH-primary care design
- Identification of the optimal strategy of various risk stratification approaches using hs-troponin testing
- Blinded, independent adjudication of clinical outcomes

*Limitations:*

- Powered for the composite MACE, but not for the individual components: death, ACS, or urgent coronary revascularization
- Despite the use of a (delayed) reference standard, complete avoidance of possible verification bias cannot be achieved

## Introduction

### Background and Rationale

Acute coronary syndrome (ACS) is a life-threatening condition requiring timely recognition and treatment to prevent severe morbidity and mortality.(3) However its clinical presentation often overlaps with far more common non-cardiac conditions such as musculoskeletal or gastrointestinal disorders (4–6). The primary diagnostic challenge is to distinguish life-threatening causes, like ACS, from more benign causes. This is particularly pronounced in healthcare systems where general practitioners (GPs) act as the 24/7 first point of contact for urgent care, yet have limited access to reliable diagnostic instruments/tools, such as cardiac troponin testing for detecting myocardial ischemia.

The hallmark symptom for ACS is chest pain, and in Dutch out-of-hours primary care (OOH-PC), GPs annually assess over 160.000 patients with chest pain (i.e. 9 per 1000 inhabitants), representing a substantial share of out-of-hours consultations, as chest pain is in the top 10 of ‘presenting complaints’(7). GP assessments rely on clinical judgement, physical examination, and, when available, an electrocardiogram (ECG). These tools may help identify alternative causes for ACS, or confirm ST-elevation, but are insufficient to safely rule out ACS (5). To minimize the risk of missed ACS, Dutch GP guidelines recommend a low referral threshold (8), and GPs generally adhere to this recommendation, resulting in high referral rates (i.e. 40-70%) for patients with chest pain to the emergency department (ED) (5, 9). This cautious approach increases burden on ambulance services, ED workload, and healthcare costs, while also causing patient distress (10). Despite high referral rates, 8-19% of ACS cases are initially missed (9, 11, 12), suggesting that the current assessment strategies in Dutch (OOH-) primary care remain suboptimal, failing to guarantee diagnostic safety while also compromising efficiency.

The development of high-sensitivity troponin (hs-troponin) point-of-care testing (POCT) may constitute a paradigm shift by bringing a rapid, user-friendly and reliable diagnostic rule-out instrument to pre-hospital care. Studies in EDs demonstrated a high diagnostic accuracy of hs-troponin POCT, showing that a single low-level troponin measurement can effectively rule-out ACS (13). However, these findings may not translate directly to primary care, where ACS prevalence is lower, diagnostic protocols are less standardized, and patient characteristics and work flows differ substantially from hospital settings.

### Objectives

To address this gap, we are conducting a diagnostic trial evaluating a primary care-tailored approach for ruling out ACS in OOH-PC that balances diagnostic safety and efficiency. Our primary objective is to evaluate the diagnostic performance of the *HEART-GP strategy*, which integrates a single fingerstick-obtained hs-troponin measurement with clinical assessment and an optional ECG in terms of safety (sensitivity and negative predictive value) and efficiency (reduction in ED referrals) when used in out-of-hours primary care. The primary outcome is major adverse cardiovascular events (MACE), a composite of death, ACS, or urgent revascularization within six weeks of index presentation, reflecting the risk for cardiovascular events rather than a single clinical syndrome.

The secondary objectives are:

1. To compare the diagnostic performance (safety and efficiency) of the *HEART-GP strategy* with current standard care based on (unaided) clinical judgment.
2. To evaluate whether using sex-specific versus universal hs-troponin cut-off values improves safety and/or efficiency.
3. To assess whether embedding hs-troponin measurements in established clinical risk scores (HEART, INTERCHEST, Marburg Heart Score) further improves diagnostic performance compared to the HEART-GP strategy alone.

## Methods

This design paper is written according to the Standard for Reporting Diagnostic Accuracy Studies (STARD) 2015 guidelines (14), and according to the Standard Protocol Items: Recommendations for Interventional Trials (SPIRIT) guidelines on defining standard protocol items for clinical trials (15).

### Study design and population

We designed a prospective diagnostic accuracy trial to evaluate the HEART-GP strategy incorporating point-of-care hs-troponin testing for ruling out MACE in patients presenting with acute chest pain or discomfort in out-of-hours primary care. The trial protocol was prospectively registered in the ISRCTN registry (ISRCTN11954040, https://doi.org/10.1186/ISRCTN11954040) and approved by the medical ethics committee of the Amsterdam UMC (NL82428.000.22).

Patient enrollment commenced in March 2023 and has been rolled out since then across four OOH-PC regions in the Netherlands: Alkmaar, Amersfoort, Leiderdorp and Venlo. Together, these regions serve approximately 1 million patients (Figure 1). Eligible patients are adults (≥18 years) who contact one of the participating OOH-PC centers regarding acute-onset chest pain or discomfort, and are triaged for a face-to-face GP consultation. Patients with signs of hemodynamic instability or chest pain following substantial trauma were excluded since both mandate immediate transfer to the ED and prohibit participation. A complete overview of the trial is displayed in Figure 2.

**Figure 1:**
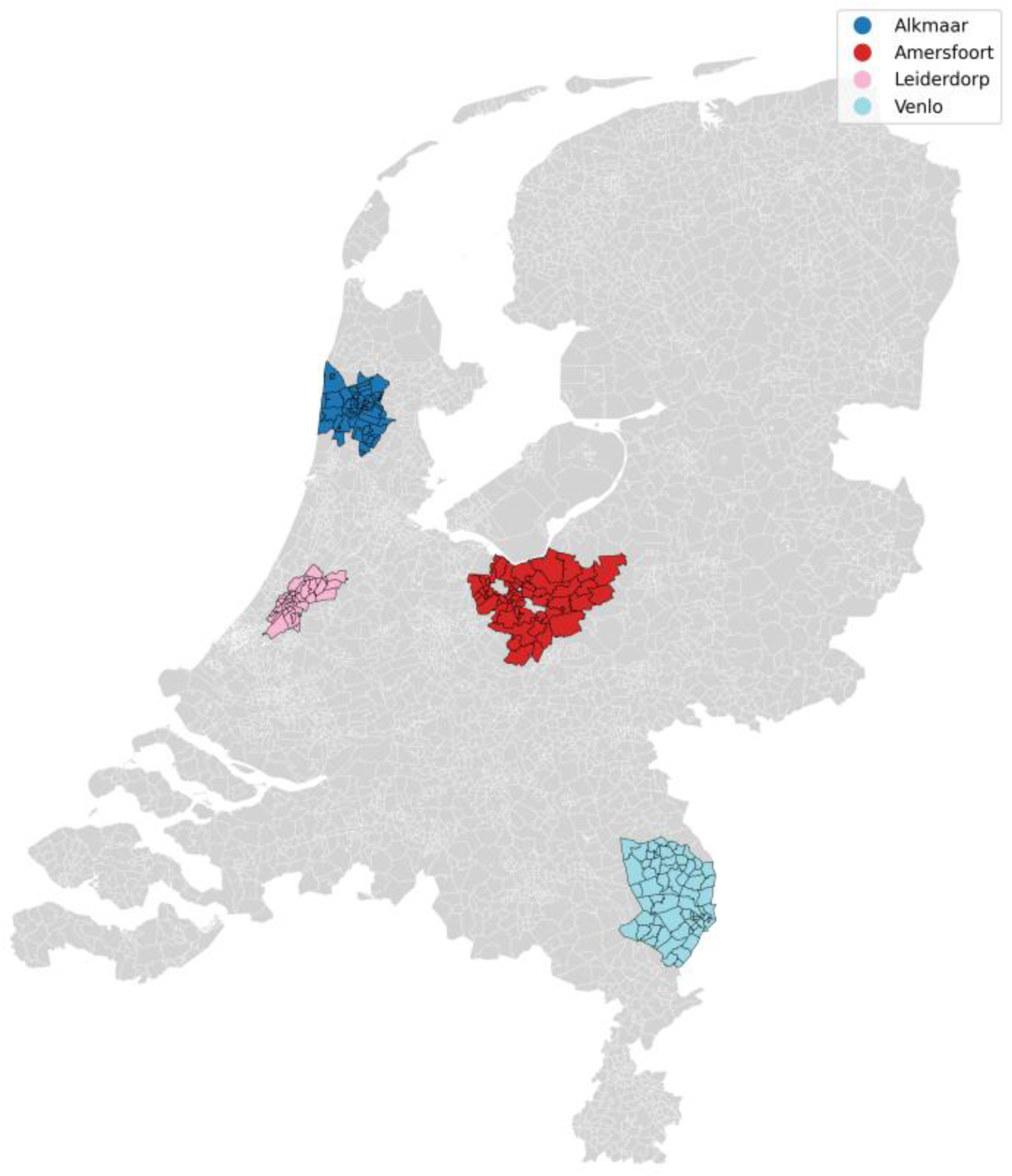
*Healthcare service area of out-of-hours primary care centers in Alkmaar/HONK* (253.000 inhabitants)*, Amersfoort/Eemland* (386.000 inhabitants)*, Leiderdorp/LIMES* (90.000 inhabitants) *and Venlo/Cohesie* (247.000 inhabitants)*, The Netherlands.*

**Figure 2:**
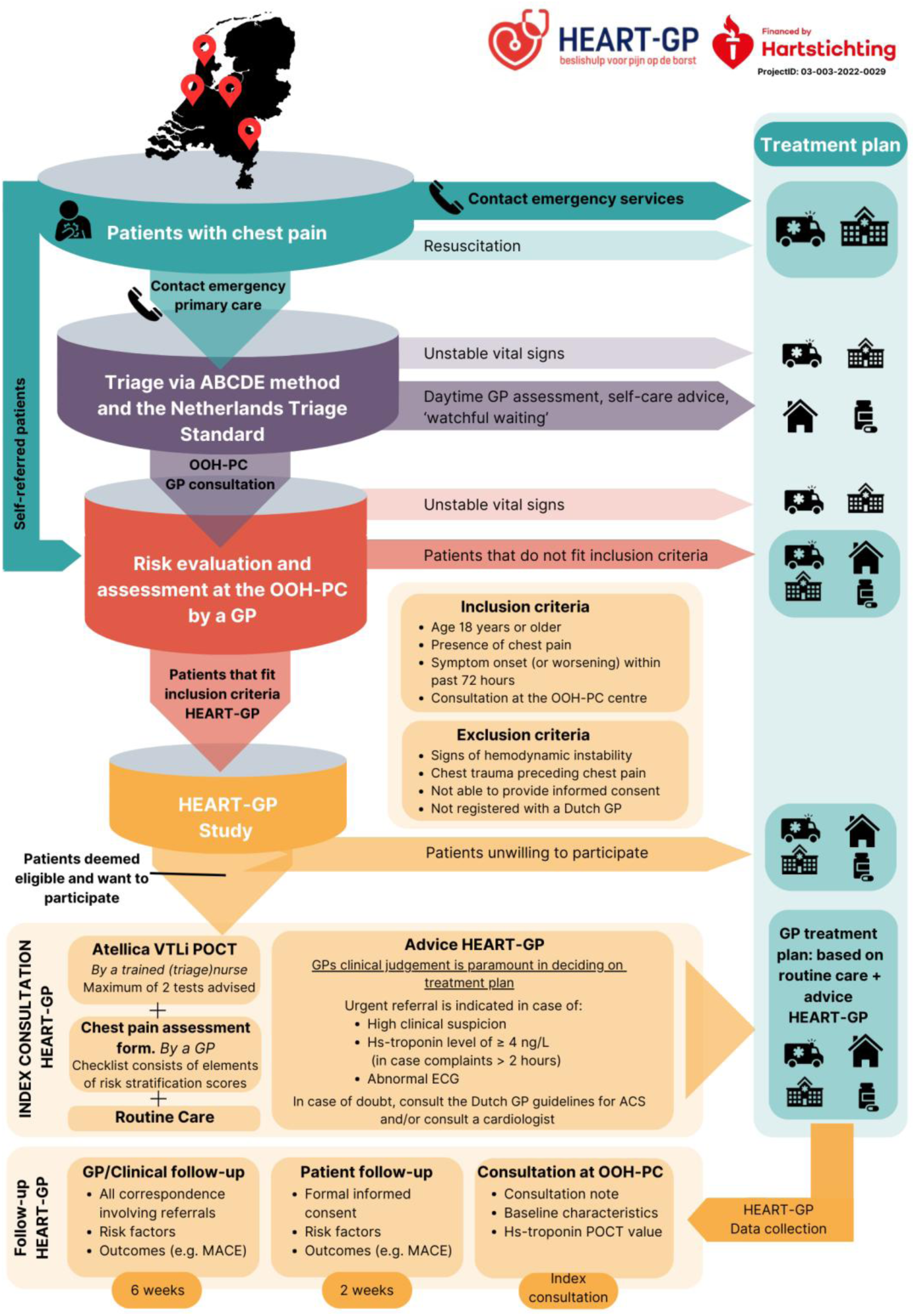
HEART-GP: patient flow, inclusion procedure and follow-up. The upper part depicts the out-of-hours workflow for patients presenting with chest pain within the Dutch healthcare system outside of office hours. The lower part (yellow) outlines the HEART-GP study workflow, including the index consultation and subsequent follow-up procedures.

##### Textbox 1: out-of-hours primary care in the Netherlands

In the Dutch healthcare system, GPs serve as round-the-clock gatekeepers to secondary care, except in emergencies requiring immediate ambulance assistance and transport to the ED. OOH-PC centers operate from 17.00 to 08.00 on weekdays and throughout weekends and holidays. They are organized through regional GP cooperatives, serving patients registered with affiliated practices within a defined geographic area (Figure 1).

Upon contact with an OOH-PC center, trained triage nurses, supervised by GPs, conduct telephone triage using the Netherlands Triage Standard (NTS), a standardized, symptom-driven protocol that assigns an urgency level to each case (1). Although the NTS provides a structured framework, triage nurses or supervising GPs may adjust the final urgency classification based on additional contextual information. This can include the patient’s perceived level of distress or whether there were repeated calls for the same symptom within a short timeframe.

Patients classified as high urgency are referred directly to the ED without GP assessment. Those with the lowest urgency level receive self-care advice, or are advised to contact their own GP during regular office hours. Patients triaged as intermediate urgency are assessed by a GP at the OOH-PC facility, where they may be treated on-site, referred to the ED for further evaluation, or discharged with appropriate safety-netting advice.(2)

### Study intervention

#### HEART-GP strategy incorporating hs-troponin testing (index test)

The intervention involves the HEART-GP strategy (Figure 3), which combines a single point-of-care high-sensitivity troponin measurement, with clinical assessment and optional ECG. We use the Siemens Atellica VTLi hs-troponin POCT to measure hs-troponin levels from a fingerstick blood sample, which returns results within 8 minutes. For ethical reasons, the GP is not blinded to the hs-troponin results during the consultation. The specifics of this assay are described elsewhere in further detail, but in essence the whole blood Limit of Detection is 1.6 ng/L and the Limit of Quantitation at 10% CV is 8.9 ng/L (16). The non-sex specific 99^th^ percentile upper reference limit for this assay is 22.9 ng/L (17). In our study we applied a lower decision threshold of ≥4 ng/L for rule out purposes, as evidence from prior studies show a sensitivity of 98.8% and a NPV of 99.8% for ruling out ACS in low-risk populations at the emergency department (ED) (13, 17).

**Figure 3:**
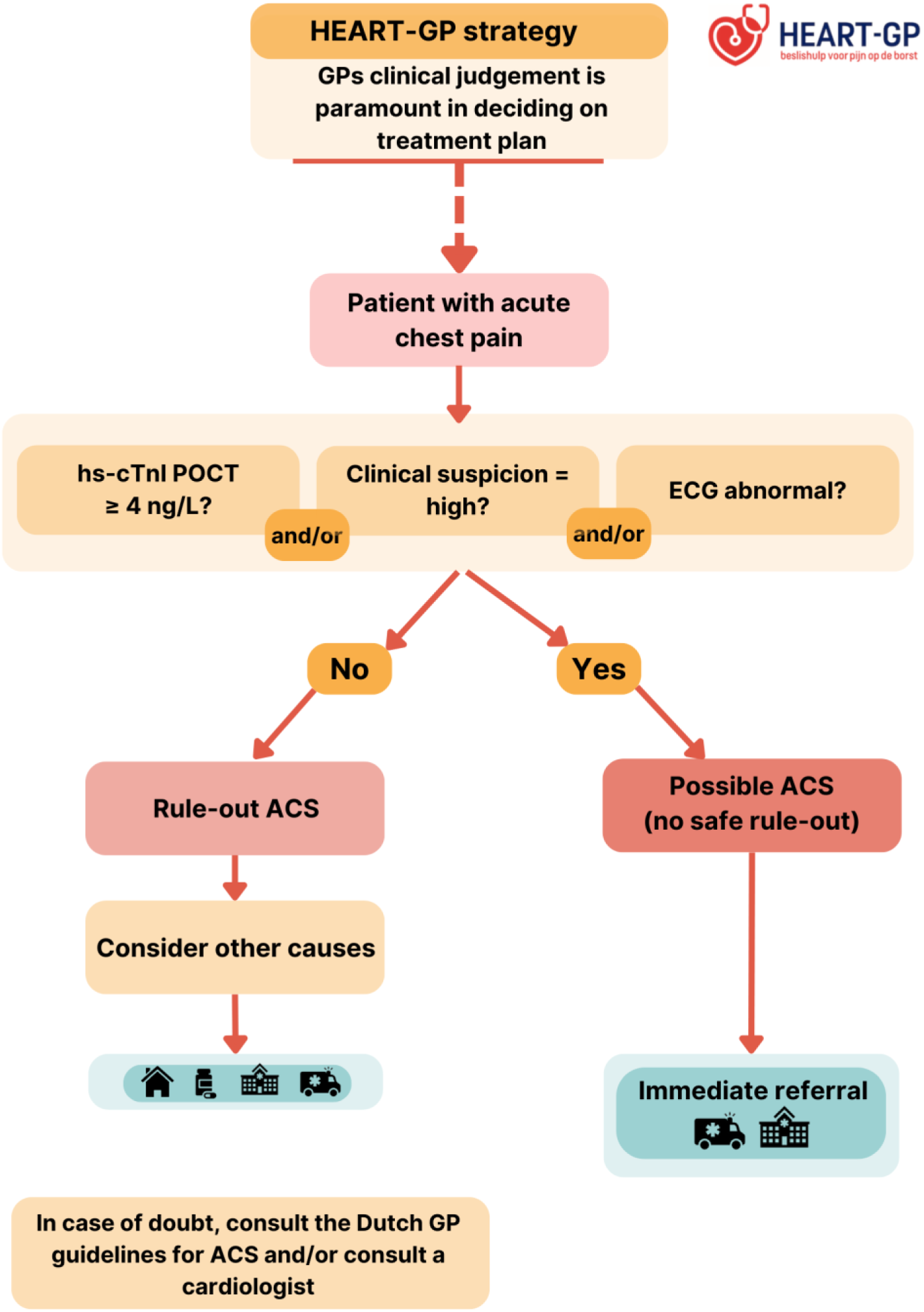
HEART-GP strategy. GP: general practitioner, hs-cTnI: high-sensitivity cardiac troponin, POCT: point-of-care test, ACS: acute coronary syndrome

All (triage) nurses at the participating OOH-PC centers receive standardized training by study personnel to ensure correct execution of the POCT, including sample collection, device operation, and interpretation of results. In the event of a test error, nurses are instructed to repeat the measurement using a maximum of two cartridges. A detailed testing protocol is included in **Appendix 1**.

The *HEART-GP strategy* guides decision-making on whether to refer the patient to the (cardiac) ED or manage them in primary care. As part of this strategy, it is of importance to note that the GPs’ clinical judgement is paramount in deciding on the treatment plan. For instance, even if a patient’s hs-troponin POCT value is below the cut-off of 4 ng/L, based on the discretion of the GP the patient may still be referred.

#### Symptom assessment form

The treating physician is asked to complete a structured, symptom-based checklist designed to standardize and document clinical assessment. The form includes symptom elements of three risk scores, namely (modified) H(E)ART, INTERCHEST and the Marburg Heart Score (Table 1). In addition, GPs are asked to record their ‘gut feeling/sense of alarm’, as well as whether they would have referred the patient based on their clinical judgement without access to the POCT result. The checklist is completed during the index consultation, instructed to be completed prior to disclosure of troponin result, to ensure unbiased responses. The form contains no recommendations or prompts regarding management decisions.

**Table 1:** (Modified) HEART(s), Marburg Heart Score, INTERCHEST risk stratification scores.

ECG: electrocardiogram, RF: risk factors for atherosclerosis, HEART: History, ECG, Age, Risk Factors, Troponin, INTERCHEST: international chest pain prediction
| (Modified) HEART score (24, 25) |  | Points | Marburg Heart Score (26) |  | Points | INTERCHEST (27) |  | Points |
| --- | --- | --- | --- | --- | --- | --- | --- | --- |
| History | Highly suspicious | 2 | Sex/age (Female ≥65, male ≥55 years) | Yes | 1 | Sex/age (Female ≥65, Male ≥55 years) | Yes | 1 |
|  | Moderately suspicious | 1 |  | No | 0 |  | No | 0 |
|  | Slightly suspicious | 0 |  |  |  |  |  |  |
| ECG | Significant ST deviation | 2 | History of cardiovascular disease | Yes | 1 | History of coronary artery disease | Yes | 1 |
|  | Non-specific repolarization abnormalities | 1 |  | No | 0 |  | No | 0 |
|  | Normal | 0 |  |  |  |  |  |  |
| Age | ≥ 65 years | 2 | Patient assumes the pain is of cardiac origin | Yes | 1 | Chest pain related to effort | Yes | 1 |
|  | 45 - 64 years | 1 |  | No | 0 |  | No | 0 |
|  | <45 years | 0 |  |  |  |  |  |  |
| Risk factors | History of atherosclerotic disease or ≥ 3 RF | 2 | Pain gets worse with exercise | Yes | 1 | Pain reproducible by palpation | Yes | Minus 1 |
|  | 1 - 2 RF | 1 |  | No | 0 |  | No | 0 |
|  | No known RF | 0 |  |  |  |  |  |  |
| Troponin* | ≥23 ng/L (99th percentile) | 2 | Pain is not reproducible by palpation | Yes | 1 | Physician initially suspected a serious condition | Yes | 1 |
|  | 4-23 ng/L | 1 |  | No | 0 |  | No | 0 |
|  | < 4 ng/L | 0 |  |  |  |  |  |  |
| *Sex-specific HEARTs: sex-specific 99<sup>th</sup> percentile for troponin (male 27 ng/L, female 18 ng/L)(17) |  |  |  |  |  | Chest discomfort feels like pressure | Yes | 1 |
|  |  |  |  |  |  |  | No | 0 |

Non-specific repolarization abnormalities: LVH: left ventricular hypertrophy, LBBB: left bundle branch block or PM: pacemaker
Risk factors: Hypertension, Hypercholesterolemia, Diabetes Mellitus, obesity (BMI >30 kg/m<sup>2</sup>), smoking, positive family history
History of atherosclerotic disease: prior Myocardial Infarction, Transient Ischemic Attack, Cerebrovascular accident, Peripheral Arterial disease or previous Percutaneous Coronary Intervention or Coronary Bypass graft

### Reference standard (occurrence of MACE)

We use a delayed-type reference test in which our primary clinical outcome is the occurrence of MACE within 6 weeks from the index consultation (see Textbox 2). MACE is a well-established endpoint accepted by regulatory agencies that captures the overall cardiovascular risk rather than focusing solely on one clinical syndrome. MACE is a composite endpoint, consisting of ACS, urgent coronary revascularization and/or all-cause death. ACS is defined as the occurrence of ST-elevation myocardial infarction (STEMI), non-ST-elevation myocardial infarction (NSTEMI), or unstable angina. Secondary outcomes are (I) the occurrence of (non)fatal myocardial infarction (STEMI or NSTEMI), (II) any hospitalization for a cardiovascular cause (e.g. heart failure, atrial fibrillation, aortic pathology, pulmonary embolism) and (III) all-cause death.

##### Textbox 2: Why we use major adverse cardiovascular events (MACE)

The primary outcome of MACE, a composite of ACS, urgent revascularization, or all-cause death/mortality at 6 weeks follow-up, is an outcome that is patient-centered, objective, and delayed, serving as a suitable reference standard in urgent primary care where not all patients undergo the same in-hospital diagnostic work-up. Using MACE rather than ACS avoids circularity with the index diagnosis and reduces verification bias. It provides a comprehensive and clinically meaningful measure of cardiovascular risk, enhances statistical power, and aligns with established regulatory and methodological standards. All events will be adjudicated with blinding to the index test, with prespecified sensitivity analyses, on the individual components of MACE, performed.

### Study flow

#### Index contact

The study workflow is described in Figures 2 and 4. In short, during the consultation the patient receives information about the study’s objectives, procedures, potential risks and benefits, and decides on their participation. Upon agreement, initial informed consent is obtained by the treating physician, and hs-troponin POCT is conducted by a trained (triage) nurse. While the POCT runs, the GP completes the ‘*chest pain assessment form*’. Subsequently, the GP determines the treatment plan ((e.g. discharge with safety-net advice (‘watchful waiting’), consulting a cardiologist or referral to the hospital for further assessment)). At all times, the GP’s own clinical judgement serves as the guiding principle in the management following the consultation. For safety reasons, urgent referral is advised in cases of high clinical suspicion of ACS, hs-troponin POCT values ≥ 4 ng/L, and/or an abnormal ECG (if performed), in line with the HEART-GP strategy recommendations. Additionally, GPs are informed to refer all patients with troponin levels exceeding the overall 99^th^ percentile of 22.9 ng/L. Patients who refuse study participation receive routine care only, which focuses on history taking, physical examination and, based on the discretion of the GP, potentially an ECG, as this is not a part of the standard work-up of chest pain in Dutch primary care (8).

**Figure 4:**
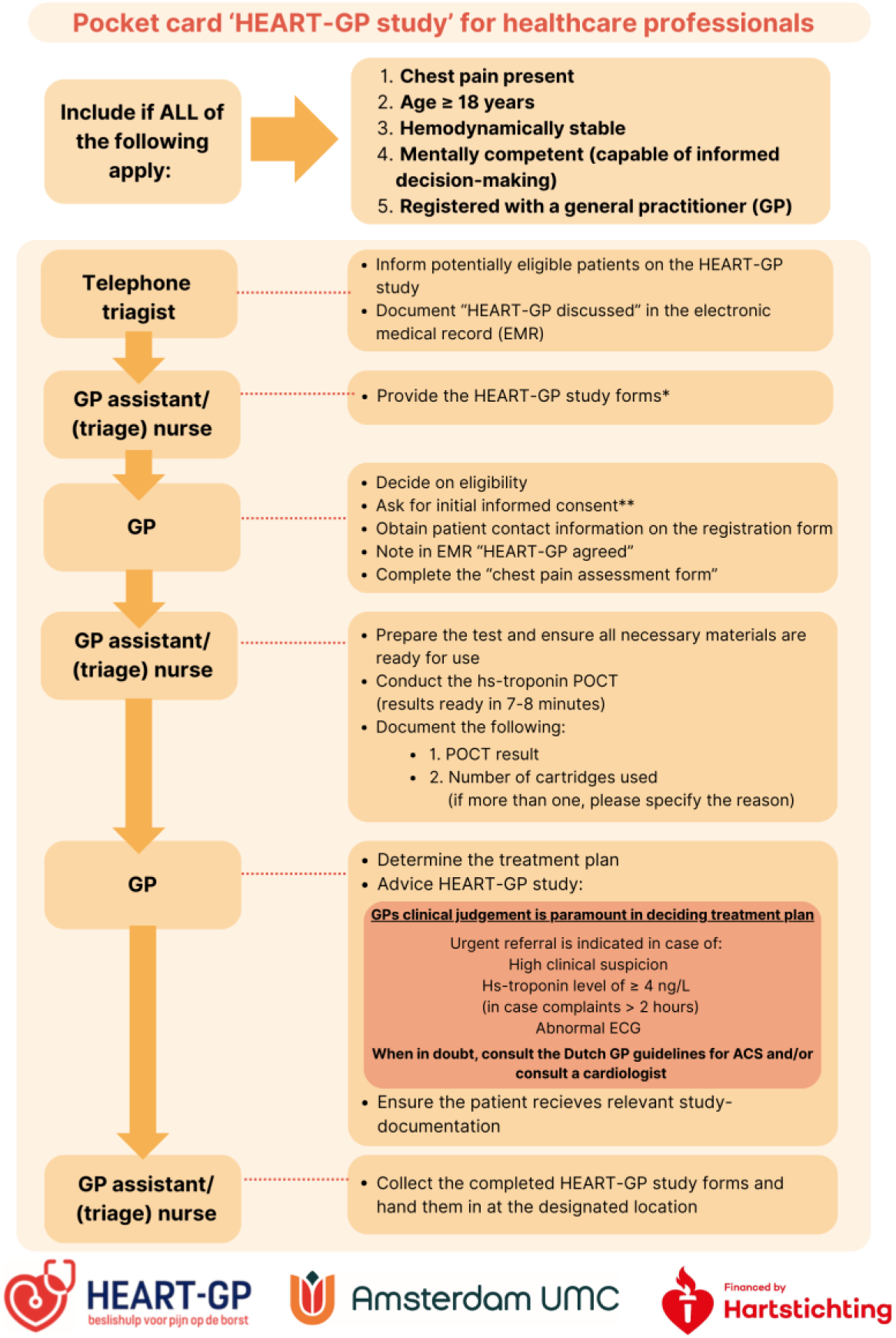
Study flow at index consultation. *HEART-GP study forms include the following: pocket card ‘HEART-GP study’ for healthcare professionals, registration form, Chest pain assessment form, Informed consent form, retour envelope, Patient Information Form (PIF) and an accompanying letter. **Initial informed consent is asked during index consultation, the patient is offered the following information beforehand. *“This out-of-hours primary care center is conducting research involving patients experiencing ‘chest pain’. As part of the HEART-GP study, you will be asked a few additional questions about your symptoms, and an additional finger-stick test will be performed. This test helps to assess whether further diagnostic evaluation at the hospital may be necessary. Please note: the test does not itself diagnose a heart attack. A result exceeding 4 ng/L, does not allow for the definitive exclusion of a heart attack. If you agree to participate, the fingerstick test will be conducted and the research team will contact you shortly. They will request your formal informed consent. You may withdraw from the study at any time without consequences. Your personal information will be treated with strict confidentiality.* *POCT: point-of-care test, ECG: electrocardiogram, ACS: acute coronary syndrome*

#### Follow-up contacts and outcome data

Two weeks after the index consultation, research staff will contact participating patients to finalize written informed consent and obtain patient-reported risk factors and outcome data. Furthermore, baseline characteristics, outcome data, and other trial data are prospectively and systematically collected for up to 6 weeks following the index consultation (Figure 2). Outcome data for all participants, including those lost to follow-up (no follow-up outcomes available), are obtained from medical records at the patients’ general practice and from regional hospital databases. All final diagnoses, including MACE, are adjudicated by an expert panel. The expert panel consists of a GP with a special interest in cardiovascular care and a cardiologist, both blinded to troponin POCT results. In case of disagreement an independent third expert will be consulted.

### Data management

Data are collected using Castor EDC (18), a secure, cloud-based electronic data capture platform with an audit trail. Data is coded, pseudonymized, and stored in accordance with General Data Protection Regulation guidelines to ensure participant confidentiality (19). To promote data quality and minimize interobserver variability, all assessors received comprehensive training on the study protocol and data entry procedures and Standard Operating Procedures (SOPs) are in place. Quality control measures such as automated range checks are employed to identify and correct discrepancies. Access to the database is restricted to authorized personnel only. Data are handled in accordance with a predefined data management plan, which includes protocols for secure storage, access, and continuous quality monitoring.

### Statistical analysis

Data will be analyzed according to the intention-to-treat principle. Baseline characteristics will be presented as numbers and percentages for categorical variables and as medians with interquartile ranges for non-normally distributed continuous variables. Categorical variables will be compared using Pearson’s chi-squared or Fisher’s exact test, and continuous variables using the Kruskal–Wallis test. The primary analysis will evaluate the diagnostic performance of the HEART-GP strategy in terms of diagnostic metrics, focusing primarily on safety, defined by sensitivity and negative predictive value for ruling out 6-week MACE, and efficiency, defined as the proportion of patients classified as low-risk. Secondary analyses will compare the diagnostic performance of the *HEART-GP strategy* with current standard care based on unaided clinical judgment, evaluate whether sex-specific hs-troponin cut-offs improve efficiency without compromising safety, and assess whether incorporating hs-troponin measurements into established clinical risk scores (HEART, INTERCHEST, and Marburg Heart Score) further improves diagnostic accuracy. Discriminatory ability for HEART-GP and comparator scores will be quantified using the area under the receiver operating characteristic curve (AUC). Differences in discriminatory performance will be tested using the DeLong method and the Net Reclassification Improvement (NRI). An exploratory analysis will examine the added value of including ECG findings in a modified HEART score. Missing data will be addressed through multiple imputation to ensure robustness of estimates.

#### Sample size calculation

Prior ED studies have reported single time-point hs-troponin sensitivities of 97-99% in patients with ≥2 hours of symptoms (13). These sensitivities are considered adequate to safely rule out ACS in primary care (20). Therefore, we applied a target sensitivity of 98% to calculate the required sample size for our diagnostic accuracy trial, comparing the index test with the reference standard (occurrence of MACE within 6 weeks of the index test). We set a margin of error of 3% around the sensitivity estimate and an alpha of 0.10 (two-sided), reflecting the rule-out focus in primary care. Assuming a MACE prevalence of 7% (literature and historic data range 5-15%), these parameters yield a required 842 participants to ensure adequate statistical power to detect a meaningful diagnostic performance in terms of sensitivity. To account for potential drop-out or a lower event rate, we aim to include at least 900 participants. Sample size calculations were performed following the methods of Akoglu, Bujang and Adnan, and calculated using PASS software (21–23).

### Safety and monitoring

To ensure patient safety, several safeguards are in place. First, hemodynamically unstable patients are excluded from participation. Second, a referral threshold of 4 ng/L is applied (well below the 99^th^ percentile) to minimize the risk of missed cases. Third, internal safety monitoring in complemented by an independent auditing monitor who conducts regular site visits to participating (OOH-PC) centers and performs random inspections of data management procedures. The monitoring body has full excess to the study data at all times. Lastly, an independent Data Safety Monitoring Board (DSMB) has been appointed to oversee patient safety, including monitoring of false negatives and serious adverse events, as well as the adequacy of participant enrollment. Enrollment is continuously monitored at each participating site. If inclusion rates decline, the study team will visit the site to identify barriers and provide support. We will also provide regular updates on recruitment progress with sites, and small incentives are offered upon reaching predefined enrollment milestones.

## Data Availability

Access to the final study dataset will be limited to the principal investigators and designated members of the research team. Any request for access to the dataset or statistical code must be approved by the study's principal investigators and will require a formal data-sharing agreement to ensure data protection and integrity.

## Statements

### Patient and public involvement statement

A representative from Harteraad, the patient advisory council for individuals with heart disease, was actively involved in the study design, protocol development, and the creation of patient information materials.

### Ancillary and post-study care

If participants experience harm related to the study, they will be referred for appropriate medical care, and compensation claims will be handled by the study’s insurance, in compliance with Dutch regulations.

### Author contributions

The study sponsor is the executive board of the Amsterdam University Medical Center. The study protocol was written by Ralf Harskamp, Simone van den Bulk and Amy Manten. This manuscript was written by Indra Melessen and Ralf Harskamp. Other authors provided feedback and revisions, all authors approved of the published version. Jos Kanning designed Figure 1.

## Acknowledgements

Lastly, we thank the OOH-PC centers (huisartsenpost Eemland, HONK, Cohesie and LIMES) and affiliated primary care practices and their staff. We would like to sincerely acknowledge and thank several staff members for their contribution at the study sites: Jack Baaij, Tirza Volk, Carla van Velden-Hollander, Natalie Fermont-Fleuren, Saine Sinnecker, Remco Rietveld, Corrie Vellema, and Liesbeth van der Plas. Lastly, we thank all the patients for participating in the HEART-GP study.

