## Appendix 1 for "The HEART-GP strategy for ruling out acute coronary syndrome in out-of-hours primary care: a diagnostic accuracy trial protocol"

### Supplement 1:

Before the administration of the test, check for initial informed consent and make preparations.

1. Make sure the participant's hand is clean and warm by holding it under warm running water or placing it in a warm bath to prevent the need for squeezing the finger.
2. Twist the cap of the lancet until it comes off, then remove it. If using a different type of lancet, select the thickest setting.
3. Let the hand hang down, press the lancet firmly against the finger, and press the button to prick.
4. After the prick, the needle automatically retracts into the lancet.
5. Remove the first drop of blood.
6. Wait patiently until the drop is large enough, then use the pipette to collect the blood without squeezing the finger.
7. Hold the pipette horizontally and touch the blood drop with the tip. Do not place your fingers near the black line, as there are air holes there.
8. The pipette fills automatically to the line and stops when full. Do not squeeze the pipette bulb while filling, and do not fill it in multiple steps.
9. Empty the pipette onto the cassette by gently squeezing the bulb.
